# Health outcomes in Deaf signing populations: a systematic review

**DOI:** 10.1101/2024.01.26.24301843

**Authors:** Katherine D Rogers, Aleix Rowlandson, James Harkness, Gemma Shields, Alys Young

## Abstract

**Objectives:** (i) To identify peer reviewed publications reporting the mental and/or physical health outcomes of Deaf adults who are sign language users and to synthesise evidence; (ii) If data available, to analyse how the health of the adult Deaf population compares to that of the general population; (iii) to evaluate the quality of evidence in the identified publications; (iv) to identify limitations of the current evidence base and suggest directions for future research.

**Design:** Systematic review.

**Data sources:** Medline, Embase, PsychINFO, and Web of Science

**Eligibility criteria for selecting studies:** The inclusion criteria were Deaf adult populations who used a signed language, all study types, including methods-focused papers which also contain results in relation to health outcomes of Deaf signing populations. Full-text articles, published in peer-review journals were searched up to 13^th^ June 2023, published in English or a signed language such as ASL (American Sign Language).

**Data extraction:** Supported by the Rayyan systematic review software, two authors independently reviewed identified publications at each screening stage (primary and secondary). A third reviewer was consulted to settle any disagreements. Comprehensive data extraction included research design, study sample, methodology, findings, and a quality assessment.

**Results:** Of the 35 included studies, the majority (25 out of 35) concerned mental health outcomes. The findings from this review highlighted the inequalities in health and mental health outcomes for Deaf signing populations in comparison with the general population, gaps in the range of conditions studied in relation to Deaf people, and the poor quality of available data.

**Conclusions:** Population sample definition and consistency of standards of reporting of health outcomes for Deaf people who use sign language should be improved. Further research on health outcomes not previously reported is needed to gain better understanding of Deaf people’s state of health.

**Strengths and limitations of this study:** This is the first systematic review of health and mental health outcomes that focused solely on evidence concerning Deaf adults who are sign language users as a distinct population rather than their incorporation within broader based health outcomes studies about deaf people in general. The review is international in scope and covers the health outcomes of Deaf adult sign language users across the globe.

This systematic review was carried out following the PRISMA guidelines by a multidisciplinary team of Deaf and hearing health service researchers from varied backgrounds.

The weakness of many studies is clearly distinguishing the population of deaf sign language users within their samples, results in discarding some evidence that might have otherwise been helpful.

**Original protocol for the study:** Prospero registration: https://www.crd.york.ac.uk/prospero/display_record.php?RecordID=182609

## INTRODUCTION

Globally, WHO estimate that 466 million individuals are living with what they define as a “disabling hearing loss” with this figure expected to reach over 700 million by 2050 [1]. Of those it is estimated that over 70 million people use one of over 300 sign languages worldwide [2, 3]. Signed languages are not visual versions of the spoken language of a country or nation, they are separate, fully grammatical living languages in their own right [4]. Deaf individuals do not perceive being deaf as a disability, and together form a community, with their own distinct language, culture and history [5]. Conventionally the upper case ‘Deaf’ is used to distinguish them from the greater population of deaf people who are spoken language users and not affiliated with Deaf communities. This distinction between deaf and Deaf is not based on degrees of deafness in an audiological sense, but is rather a sociological distinction, based on cultural-linguistic identity.

Poorer health and mental health outcomes among Deaf communities have been previously observed when compared with the general population [6, 7, 8, 9]. Suboptimal management of physical health conditions is also common, posing not just immediate health risk but increasing the risk of long-term complications. A UK study using the EQ-5D-5L recorded a mean health-state value of 0.78 for Deaf people compared to the mean health-state value for the general population of a similar age of 0.84 [10]. Common mental health problems have been found to be more prevalent amongst Deaf people in comparison with the hearing population [6]. Additionally, Deaf people are more likely to be victims of physical, sexual, and emotional abuse along with neglect, all of which are significant risk factors for poor mental health [11, 12]. Wide-spread difficulties in accessing health-related information in a signed language, accessing health care in a timely manner, cultural-linguistic barriers in interactions with clinicians and health care providers, and inappropriate diagnostic assessments normed on hearing populations have all been recorded as potential contributors to poorer health outcomes in this population [13, 7, 14].

However, a comprehensive systematic review of the evidence concerning the physical and mental health outcomes of signing Deaf adult populations has yet to be undertaken. PROSPERO records a current systematic review of Inequities Experienced by Deaf and Hard of Hearing Patients in Healthcare Access and Healthcare Delivery [CRD42020161691] and one concerning the prevalence and correlates of mental and neurodevelopmental symptoms and disorders among deaf children and adolescents [CRD42020189403]. Neither addresses health and mental health outcomes of Deaf signing adults which is required as a guide to future research and to assist clinicians in their current work.

### Research questions

- What does the available literature conclude about the mental and physical health of adult Deaf population(s)?

`o` How does the health of the Deaf population(s) compare to that of the general population(s)?
`o` What are the strengths and weaknesses of the available literature?
`o` What should future research aim to address?

## METHODS

### Original protocol for the study

Prospero registration: https://www.crd.york.ac.uk/prospero/display_record.php?RecordID=182609

### Eligibility criteria

An electronic literature search was used to identify relevant studies up to (13^th^ June 2023). No starting cut-off date was applied. The following electronic databases were searched using the OVID platform: Medline; Embase; PsychINFO; and Web of Science. The research strategy included the keywords (e.g. deaf*, health*, and sign*) and the key terms were truncated and combined through use of the Boolean operators ‘AND’ and ‘OR’ (see online supplementary table S1).

Articles identified by the search underwent a two-step screening process. Each stage of the review was completed independently by two reviewers ([AUTHOR ONE] & [AUTHOR TWO]), with a third reviewer ([AUTHOR FIVE]) consulted to settle any disagreements. Firstly, publication titles and abstracts were screened for relevance using the inclusion and exclusion criteria, defined prior to searching (see Table 1). The papers that met these criteria were then screened by a full paper review, again using the predefined eligibility criteria.

**Table 1.** Inclusion and exclusion criteria.

| Aspect of study | Inclusion | Exclusion |
| --- | --- | --- |
| Population | Signing Deaf populations*<br>Adults (aged 18+)<br>Studies focusing on a subgroup of Deaf populations (e.g. LGBTQ, those with learning disabilities/difficulties) | deaf populations (with a lowercase 'd') and/or where deaf is not used to define those who are sign language users<br>Hard-of-hearing populations who are not sign language users<br>Children or adolescents<br>Those with single sided deafness<br>Individuals who are blind/ those with dual sensory impairment<br>Individuals with age-related hearing loss<br>Cochlear implant users who are not sign language users<br>Deaf people who use spoken language exclusively |
| Study type | All study types other than those in the exclusion criteria<br>Full text<br>Peer-reviewed<br>Primary or secondary peer reviewed research articles | Letters<br>Editorial/opinion pieces<br>Historical articles<br>Case reports<br>Conference abstracts |
| Outcomes | Measures of mental health such as the prevalence of mental health conditions and measures used in relation to mental health (e.g., the PHQ-9 and GAD-7) | Papers reporting on the physical or mental health of carers/relatives of Deaf individuals<br>Papers reporting on hearing health conditions including their aetiology and treatment<br>Papers reporting studies on audiology |
|  | Measures of physical health, such as the prevalence of chronic conditions and symptom measures for physical health conditions<br>Measures of overall health status, quality of life and wellbeing, that may reflect mental and physical health combined<br>Morbidity, mortality and prevalence statistics | Papers reporting on correction or improvement to hearing<br>Papers reporting on health measurement instruments (such as questionnaire validation studies) without data on health status.<br>Papers reporting on barriers to health care or health delivery to Deaf populations that exclude any health outcome data<br>Papers reporting on health risk behaviours unless containing data on specific health outcomes |
| Country | No restriction by country |  |
| Language | Research articles not published in English language nor a signed language |  |
| Timeframe | Up until 13<sup>th</sup> June 2023 |  |
| * Signing Deaf populations are defined using a sociological/cultural-linguistic definition, referring to a particular group, not defined by the audiological condition of not hearing, but rather those who use a signed language as their first or preferred language— and share a culture. There is not one global population but signing Deaf populations exist in each country.<br>Abbreviations: GAD-7, Generalised Anxiety Disorder assessment-7; PHQ-9, Patient Health Questinnaire-9; LGBTQ, Lesbian Gay Bisexual Transgender and Queer. |  |  |

### Data extraction

Comprehensive data extraction was performed using a pre-specified data extraction tool based on the Cochrane data collection form (<u>Collecting data - form for RCTs and non-RCTs</u>) and additional data extraction criteria to accommodate a range of study designs. This included extracting information on study samples, methodology, limitations, evidence gaps, results, and a quality assessment for critical appraisal. Three reviewers ([AUTHORS ONE, TWO, and THREE]) extracted data independently. Results were then compared and discussed, with any disagreements settled between them and an additional reviewer ([AUTHOR FIVE]).

### Quality assessment

The CASP Cohort Study checklist (https://casp-uk.net/casp-tools-checklists/) was used to assess bias in each study as it was more appropriate to the range of items than other more design-specific checklists in the CASP suite. Two reviewers ([AUTHORS ONE AND TWO]) independently assessed the risk of bias, with quality checks performed on 25% of the extracted papers by a third reviewer ([AUTHOR FIVE]), to ensure consistency. Results were compared with any disagreements resolved by a third reviewer.

## RESULTS

Screening outcomes are presented in Figure 1.

**Figure 1.**
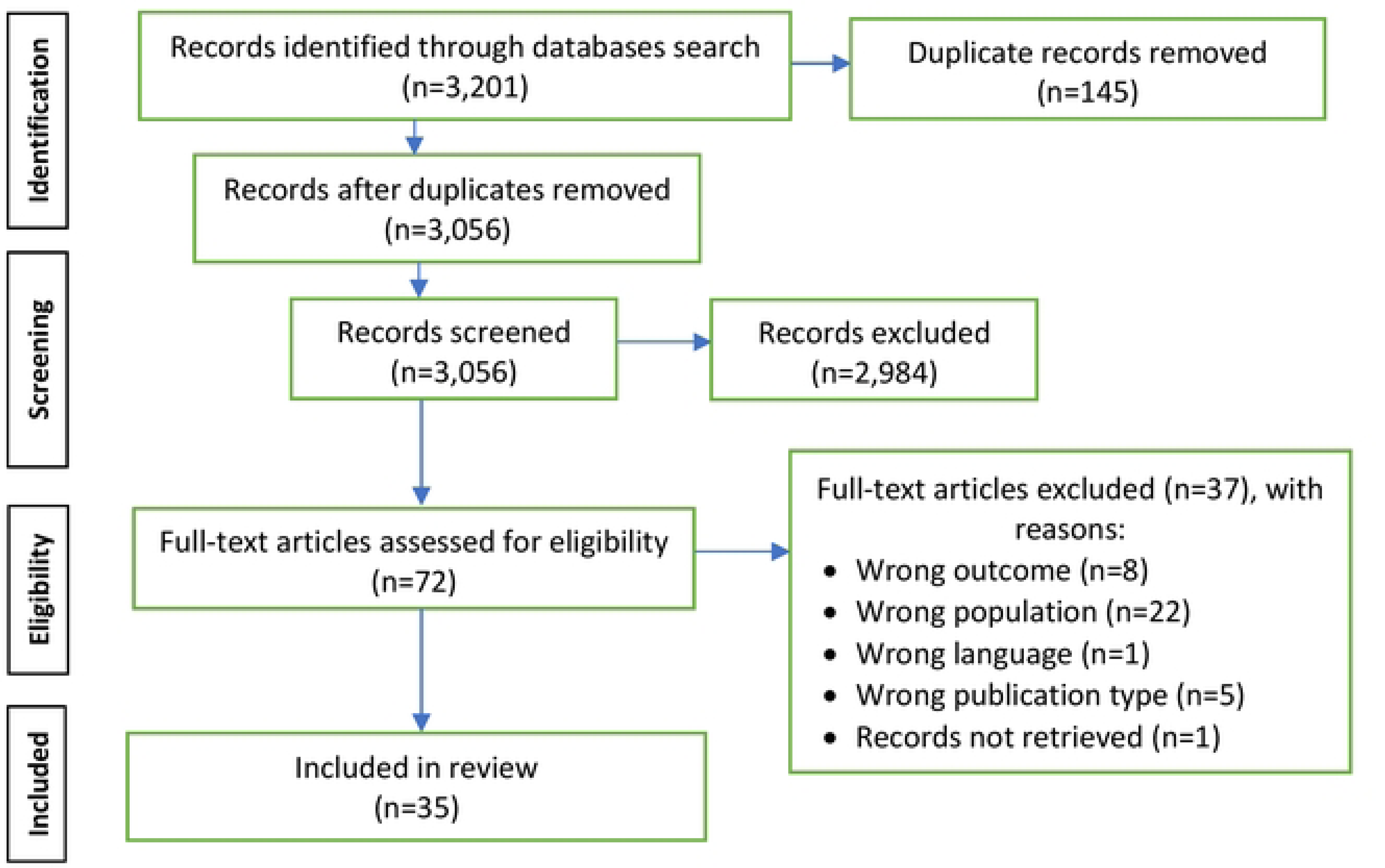
**PRISMA flow diagram: the findings from the searches**

### Study characteristics

All 35 items included were peer review journal articles of which 27 reported primary data and 8 were secondary data analyses. 29 studies utilised self-report health data. The identified publications varied by research setting and sample size. 32 out of the 35 studies took place in economically well resourced, developed countries, 2 from low- and middle-income countries and 1 from least developed countries. Only one study involved a randomised controlled trial.

Of the 35 studies, 26 used comparator populations in their study designs. Of these, 19 used comparisons with the “general population”. This was not always distinguished to mean the hearing population. For example, the general population could include people who had a hearing loss but were not signing Deaf people. Comparators were either general population reference data for a particular disease, or general population survey study samples (e.g. Health Survey of England data). Three studies [15, 16, 17] used external datasets from the general population to construct a comparison group that were to some degree matched for a range of demographic variables (e.g. age and gender). Eight studies sought comparative data between d/Deaf populations. For example, Deaf sign language users versus other deaf people who used spoken language or within Deaf communities whereby Deaf people were distinguished by intersecting characteristics such as ethnicity or sexuality.

### Study appraisal

Quality appraisal by study is shown in Table 2. Application of the two initial screening questions in CASP resulted in 28 of 35 studies being eligible for quality appraisal with tool. No studies met all the CASP criteria. Comments on the strengths/weaknesses of studies are incorporated in the presentation of health outcome data below.

**Table 2.** CASP study appraisal for each included in the review. Y = yes, CT = Can’t tell, N = no. Note: CASP criterion 7 reports the results and is excluded here as they are included in the main text. In the CASP criterion 8 (precise) Y indicates Confidence Intervals (CIs) were reported, and N that they were not.

| Author(s) (year) | CASP criterion |  |  |  |  |  |  |  |  |  |  |  |  |
| --- | --- | --- | --- | --- | --- | --- | --- | --- | --- | --- | --- | --- | --- |
|  | 1 | 2 | 3 | 4 | 5(a) | 5(b) | 6(a) | 6(b) | 8<br>(precise) | 9 | 10 | 11 | 12 |
| Ammons et al (2020) [37] | Y | CT | Y | Y | Y | Y | Y | Y | N | CT | CT | Y | Y |
| Anderson et al (2021) [33] | Y | Y | Y | Y | CT | Y | Y | Y | N | CT | CT | CT | Y |
| Barnett et al (2011) [9] | Y | Y | Y | Y | Y | CT | CT | CT | Y | Y | N | Y | Y |
| Barnett et al (2016) [28] | Y | Y | Y | Y | Y | N | CT | Y | Y | CT | N | CT | Y |
| Barnett et al (2023) [44] | Y | Y | Y | Y | Y | Y | Y | Y | Y | Y | CT | Y | Y |
| Belk et al (2016) [43] | Y | Y | Y | Y | Y | N | Y | Y | N | CT | Y | CT | Y |
| Chapman et al (2017) [35] | Y | Y | Y | CT | Y | Y | Y | Y | N | Y | Y | CT | Y |
| Crowe et al (2016) [59] | Y | CT | CT | CT | CT | N | CT | Y | N | CT | N | CT | CT |
| Druel et al (2018) [18] | Y | Y | Y | Y | CT | Y | Y | Y | N | Y | Y | Y | Y |
| Duarte et al (2021) [38] | Y | Y | Y | Y | Y | Y | CT | Y | Y | Y | Y | Y | Y |
| Ehn et al (2018) [15] | Y | Y | Y | Y | CT | CT | Y | Y | Y | Y | Y | Y | Y |
| Emond et al (2015) [8] | Y | Y | Y | Y | CT | CT | Y | Y | N | Y | Y | Y | N |
| Fellinger et al (2005) [30] | Y | Y | N | CT | Y | CT | Y | Y | N | N | CT | Y | Y |
| Henning et al (2011) [31] | Y | CT | CT | Y | Y | Y | Y | Y | N | Y | CT | Y | Y |
| Horton (2010) [60] | Y | CT | Y | Y | Y | Y | Y | Y | N | Y | CT | Y | Y |
| James et al (2022) [19] | Y | Y | Y | Y | Y | CT | Y | Y | Y | CT | CT | Y | Y |
| James et al (2022) [25] | Y | Y | Y | CT | Y | Y | Y | Y | Y | CT | Y | Y | Y |
| Kushalnagar et al (2019) [32] | Y | Y | CT | CT | Y | Y | Y | Y | Y | Y | Y | CT | Y |
| Kushalnagar et al (2019) [20] | Y | Y | Y | Y | Y | Y | Y | Y | Y | Y | Y | CT | Y |
| Kushalnager et al (2020) [39] | Y | Y | Y | Y | Y | Y | Y | Y | Y | Y | Y | CT | Y |
| Kvam et al (2007) [21] | Y | Y | Y | CT | Y | Y | Y | Y | Y | CT | Y | Y | CT |
| McKee et al (2014) [36] | Y | Y | Y | CT | Y | Y | Y | Y | Y | Y | CT | Y | Y |
| Munro et al (2009) [41] | Y | CT | Y | CT | N | N | Y | Y | Y | Y | CT | Y | Y |
| Øhre et al (2017) [40] | Y | Y | Y | CT | Y | N | CT | CT | N | CT | CT | Y | Y |
| Peñacoba et al (2020) [16] | Y | Y | Y | N | Y | Y | Y | Y | Y | Y | Y | Y | Y |
| Perrodin-Njoku et al (2022) [17] | Y | Y | Y | CT | Y | CT | Y | Y | Y | Y | Y | CT | Y |
| Rogers et al (2013) [42] | Y | CT | Y | Y | Y | N | Y | Y | N | Y | CT | CT | Y |
| Rogers et al (2016) [29] | Y | Y | Y | Y | Y | N | Y | Y | Y | Y | N | Y | Y |
| Rogers et al (2018) [23] | Y | Y | Y | Y | Y | N | Y | Y | Y | Y | N | CT | Y |
| Sanfacon et al (2021) [34] | Y | Y | Y | Y | Y | Y | Y | Y | Y | Y | Y | CT | Y |
| Shields et al (2020) [10] | Y | Y | Y | Y | Y | Y | Y | Y | Y | Y | CT | Y | Y |
| Simons et al (2018) [24] | Y | Y | Y | Y | Y | Y | Y | Y | Y | Y | Y | N | Y |
| Vichayanrat et al (2014) [26] | Y | CT | Y | CT | Y | Y | Y | Y | N | CT | CT | N | Y |
| Wahlqvist et al (2016) [27] | Y | Y | Y | CT | Y | Y | Y | Y | Y | CT | Y | Y | CT |
| Werngren-Elgström et al (2003) [22] | Y | Y | Y | Y | CT | N | Y | Y | N | Y | CT | CT | Y |

### Health conditions studied

The coverage of health conditions represented in the included articles is described according to the International Classification of Diseases 11^th^ Revision (ICD-11) with some studies encompassing more than one of the 26 main categories. 15 out of the 26 classifications are encompassed within the 35 included studies. The greatest number of studies (25 out of 35) concern ICD-11 Code 6 Mental, Behavioural or Neurodevelopmental disorders. This includes anxiety/depression, mental well-being, psychiatric disorder, mental distress, and schizophrenia (See Table 3). The remaining 11 classifications where there are no outcome studies identified are: 01 Certain infectious or parasitic; 03 Diseases of the blood or blood-forming organs; 04 Diseases of the immune system; 08 Diseases of the nervous system; 09 Diseases of the visual system; 10 Diseases of the ear or mastoid process; 14 Diseases of the skin; 17 Conditions related to sexual health; 19 Certain conditions originating in the perinatal period; 25 Codes for special purposes; and 26 Supplementary Chapter Traditional Medicine Conditions - Module I.

**Table 3.**
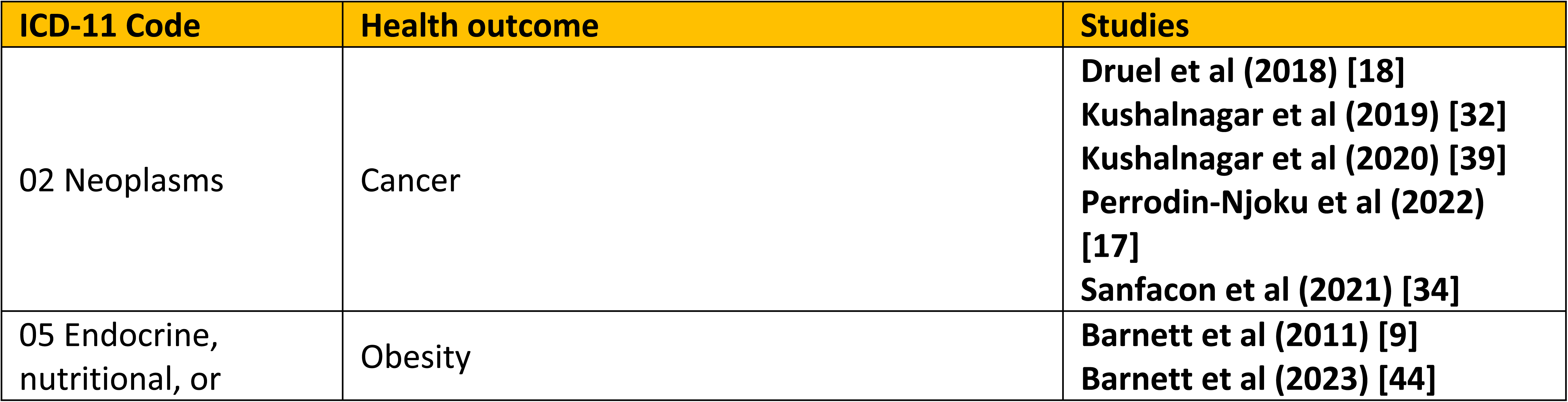
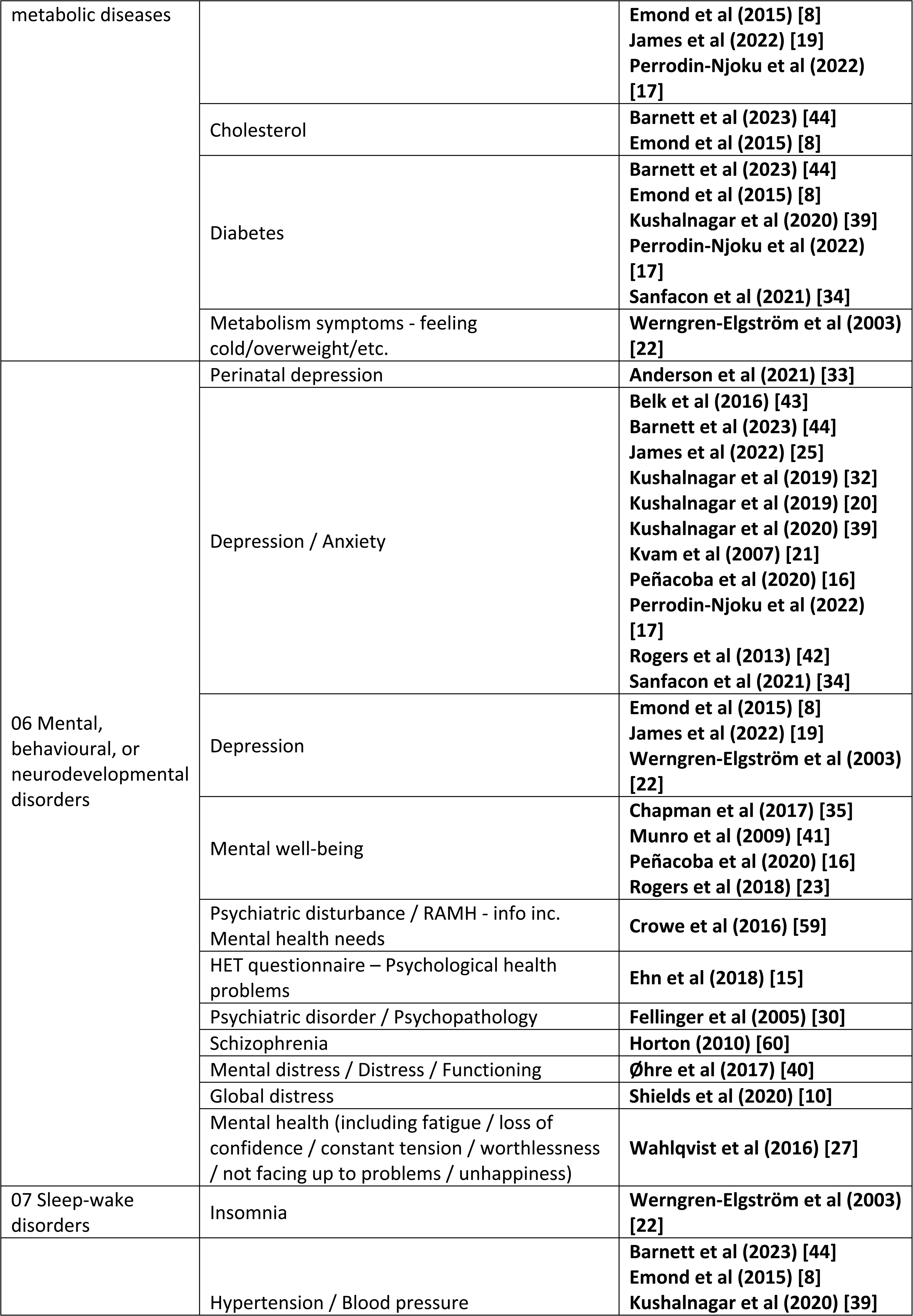
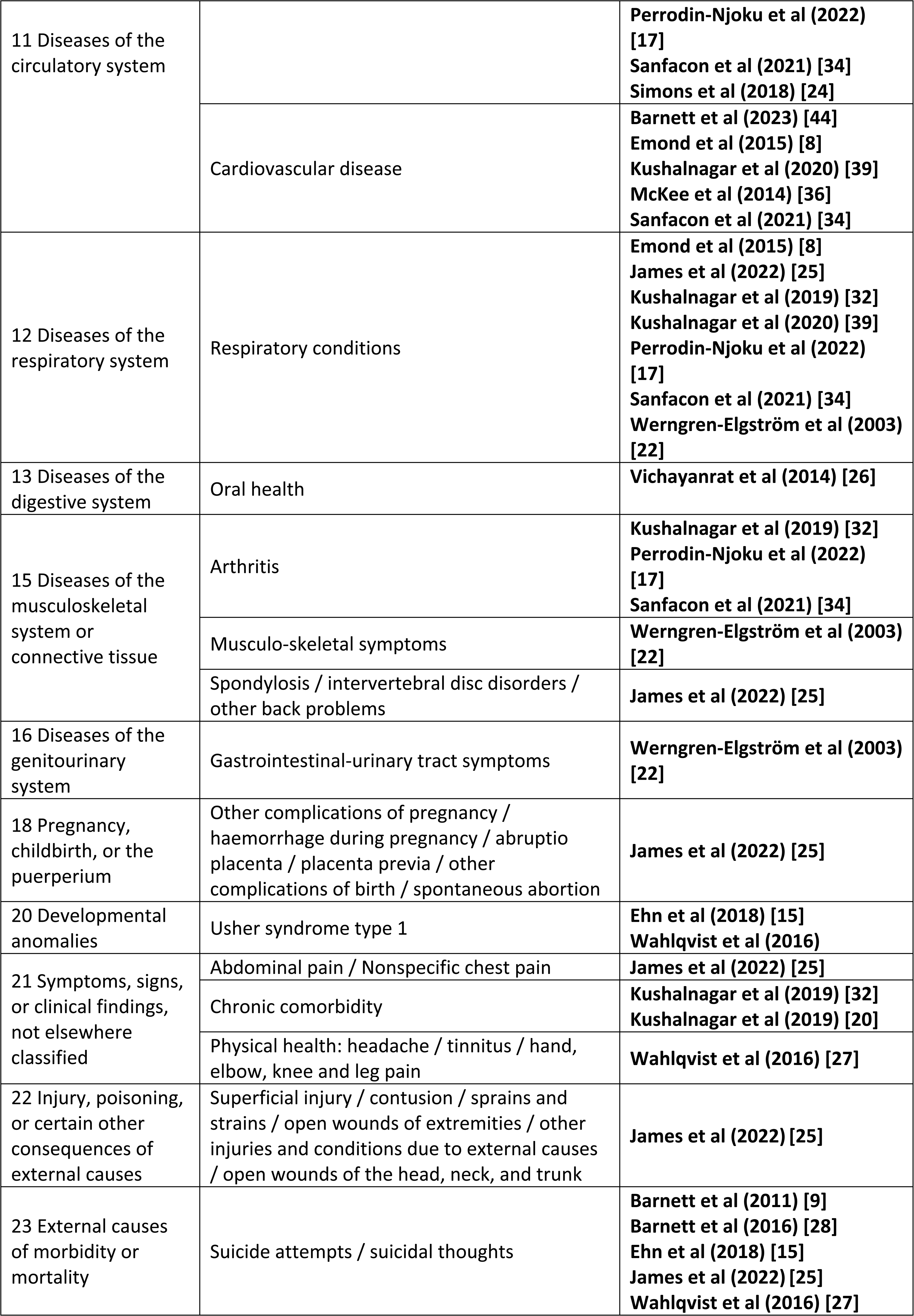
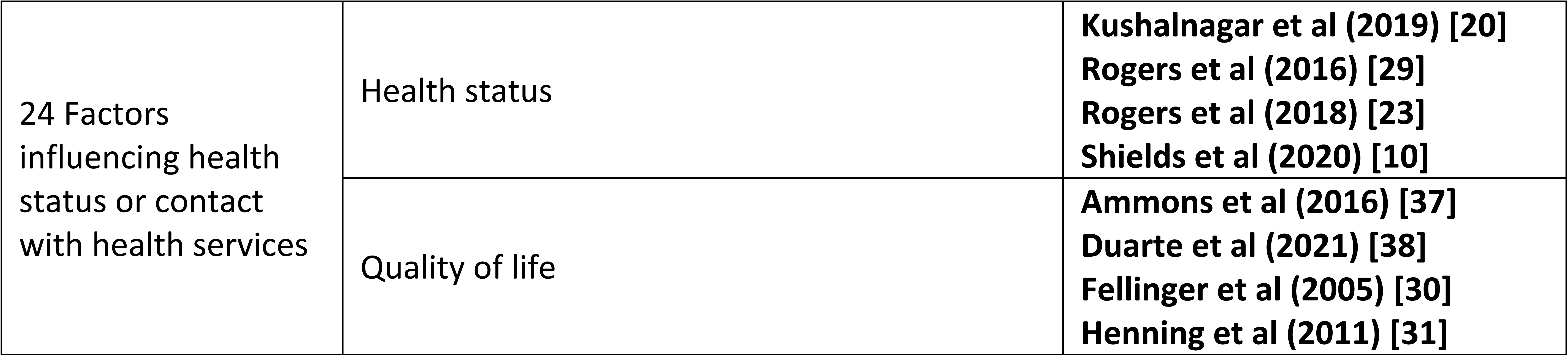
Reported health outcomes in the included studies by ICD-11 main classification code.

### Health Outcomes in comparison with hearing/general populations

**Table 4.**
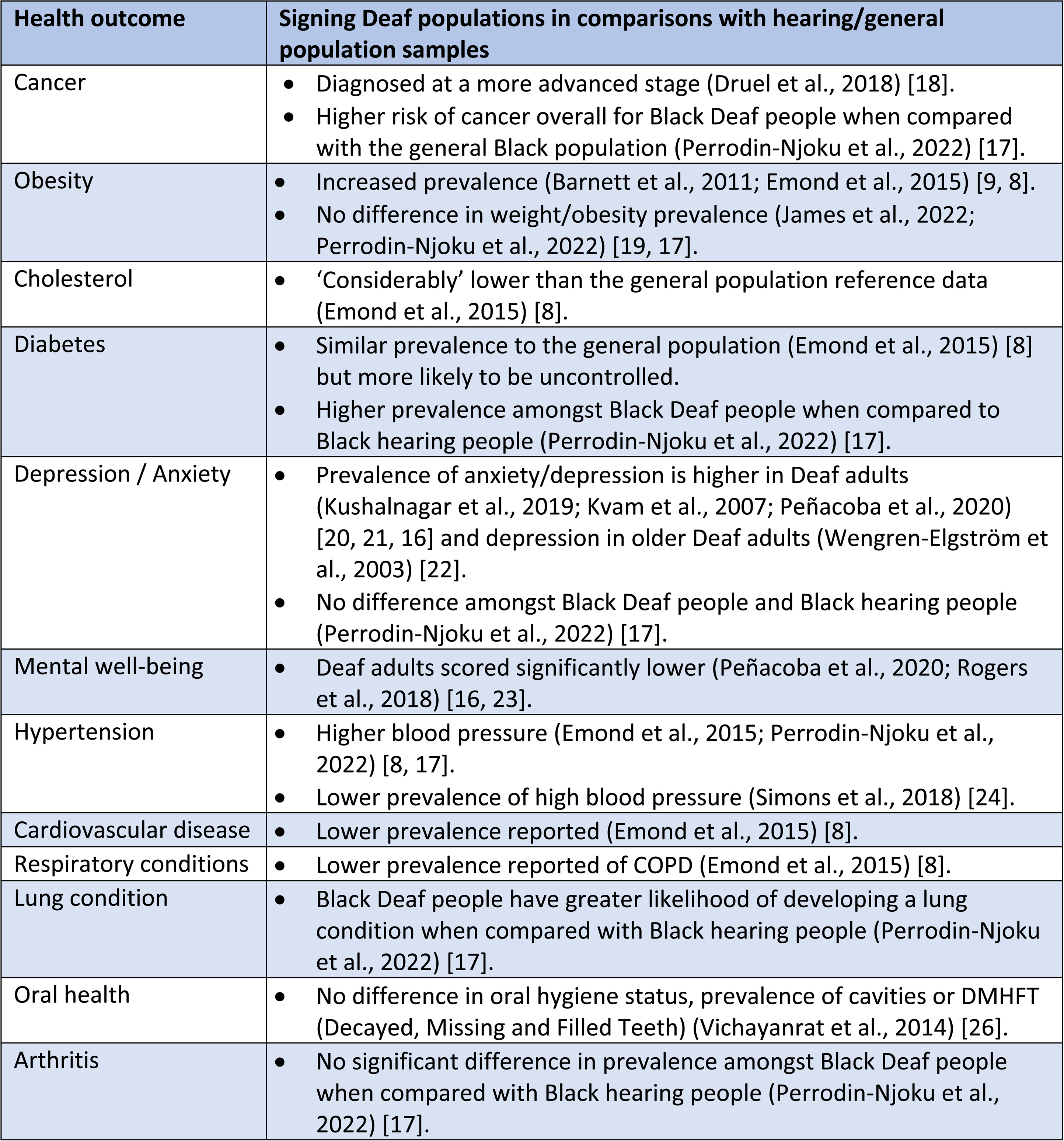
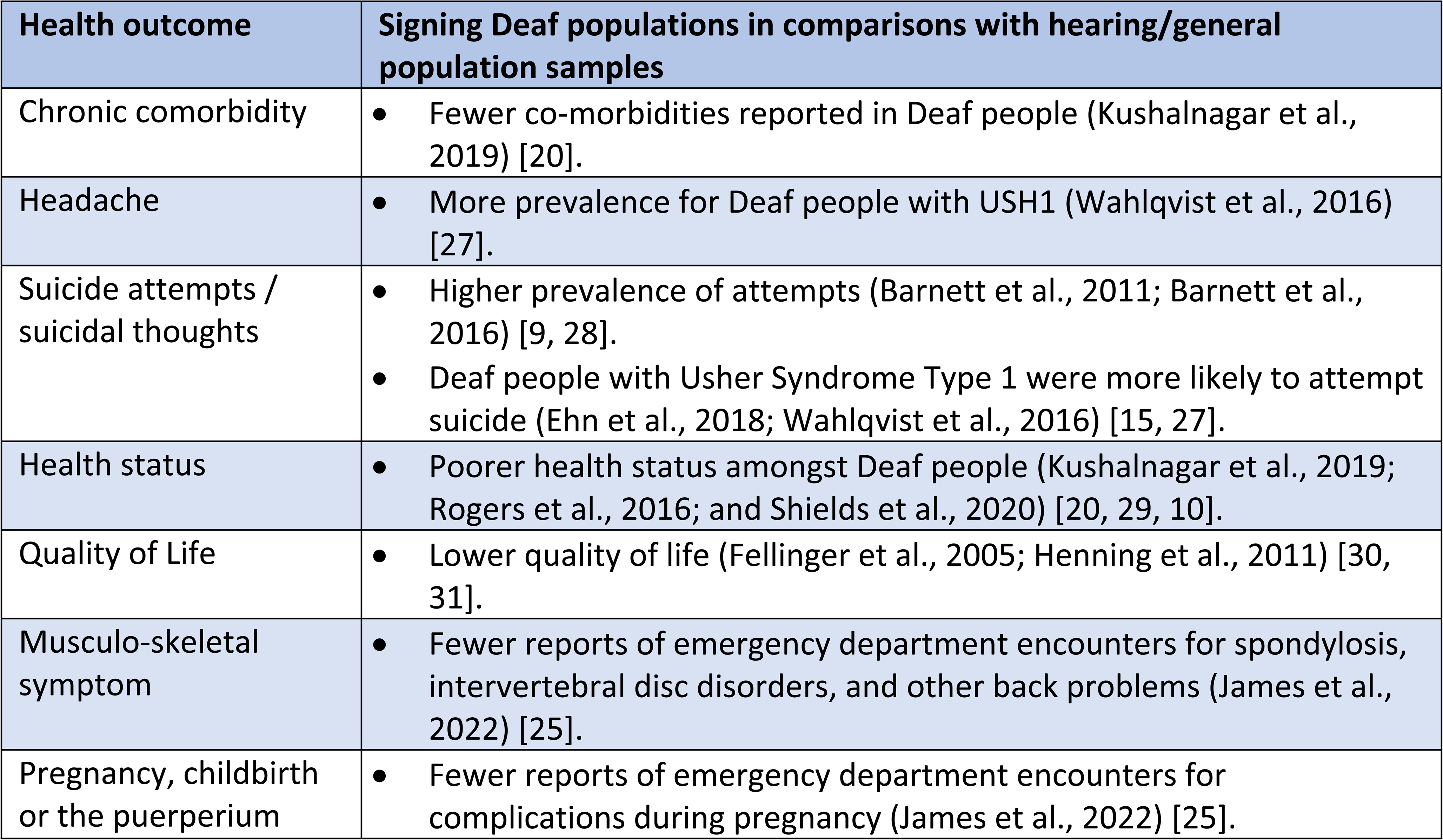
Overview of health outcomes in comparison with hearing / general populations.

### Cancer

A large-scale study [18] involving secondary data from medical records of Deaf cancer patients found that Deaf people were diagnosed at a more advanced stage of colorectal and prostate cancer (64% of the Deaf group vs 13% of the reference group for prostate cancer, and 100% of the Deaf people were diagnosed at stage III/IV vs 47% for the reference group for colorectal cancer). The Deaf group had larger tumours at the time of diagnosis and their cancers were more likely to have spread to lymph nodes or metastasised to other organs. Deaf people were also more likely to be diagnosed with larger tumours in breast cancers (T2+ size was 60% for Deaf people compared to the reference group 34%) which is related to poorer prognosis, although there was no difference in the metastatic spread between the groups [18]. In an age-matched large-scale study, Perrodin-Njoku et al [17] found that Black Deaf people were more likely to have cancer overall compared to Black hearing people (OR=3.53, CI 1.61-7.71).

#### Obesity

Two primary studies [9, 8] recruiting 339 and 298 respectively reported significantly higher rates of overweight/obesity among Deaf populations when compared to published data on general populations: 35% of Deaf adults vs 26.6% of adults in the US general population [9] and 72% of Deaf men and 71% of Deaf women were overweight or obese vs 65% of men and 58% of women in the general population in the UK [8] HSE dataset. In the Emond et al. study [8], 90% of the over 65 Deaf group were overweight or obese. Neither a small-scale patient record study (n=92) [19] nor a case-matched comparator study in Black populations [17] reported any significant differences. However, differences in the mean age of the comparator groups was noted for both studies which could help to explain the non-significant findings.

#### Cholestero**l**

Emond et al. [8] found that the mean level for both males and females was ‘considerably’ lower compared to general population Health Survey of England (HSE) reference data [8] although it was not clear whether the reported difference is statistically significant or not. No potential co-variates were examined.

#### Diabetes

Edmond et al. [8] found that of those Deaf people who reported diabetes, at least half of the participants’ diabetes was not under control, which could lead to higher rates of diabetic complications. Perrodin-Njoku et al. [17] reported that Black Deaf people are more likely to have diabetes compared to Black hearing people in the US (OR=1.77, CI = 1.04-3.02). The type of diabetes was not reported in either study.

#### Depression / Anxiety

The prevalence of depression / anxiety in Deaf adults was found to be significantly higher compared with the hearing population. A large-scale study (n=1,704) by Kushalnagar et al. [20] reported 24.9% compared with 21.7% and that it occurred at an earlier age; Kvam et al.’ Norwegian population study [21] reported 33.8% compared with 6.8%; Peñacoba et al. [16] reported mean scores for anxiety of 8.06 compared with 6.60 and for depression 5.01 compared with 3.27 in a case-matched study of Spanish Deaf and hearing adults. Perodddin-Njoku et al [17] reported no difference in the Black Deaf population compared to the Black hearing population, they stated that the issue of medical mistrust in the general Black community might be a factor in finding no difference between the two groups. Werngren-Elgström et al. [22] in a small Swedish comparative study (n=45) found that Deaf people aged 65 and over have a higher prevalence of depression compared to their hearing counterparts (37% vs 23%).

#### Mental well-being

A case-matched study of Spanish Deaf adults (n=146) reported significantly lower psychological well-being compared to Spanish hearing people: mean score of 24.58 vs 27.44 [16]. Rogers et al. [23] small scale validation study reported a non-significant lower well-being mean score on the SWEMWBS (22.82) in comparison with the general population (23.64).

#### Hypertension

The frequency of raised blood pressure was significantly higher for Deaf people (37%) compared to the HSE data (21%) although the confidence interval was not reported [8]. Perrodin-Njoku et al. [17] also reported that Black Deaf people are more likely to experience higher blood pressure compared to the Black hearing population (OR=1.73). However, a large-scale study (n=532) by Simons et al. [24] reported that the prevalence for hypertension was significantly lower in the Deaf sample (33%) compared with 46% in the hearing sample.

#### Cardiovascular disease

Cardiovascular disease was significantly less self-reported by Deaf people compared to the general population [8]. Emond et al. [8] found that treatment rate for Deaf men of all CVD was 45% compared with treatment rate for ischaemic heart disease and stroke of between 61% and 70% for men age aged 55-84 in the general population.

#### Respiratory / lung conditions

Self-reported Chronic Obstructive Pulmonary Disease (COPD) was less in Deaf adults (1%) compared to the HSE data (4% of men and 5% of women) [8] although the significant difference was not stated. Black Deaf people have a greater likelihood of developing a lung condition compared with Black hearing people (OR=1.72) [17]. Fewer DHH ASL users were reported in emergency department encounters for lower respiratory disease compared to DHH English speakers and hearing English speakers (n=11 vs n=62 and n=29 respectively) [25].

#### Oral health

Vichayanrat et al. [26] reported no differences between Deaf and hearing people in prevalence of caries or DMFT (Decayed, Missing or Filled Teeth), and similar oral hygiene status. Those Deaf people who took part in Vichayanrat et al. [26] study were educated at BA and/or Diploma level, therefore, unlikely to be representative of the Deaf population.

#### Arthritis

A self-report study by Perrodin-Njoku et al. [17] found no significant difference in prevalence of arthritis between Black Deaf people and Black hearing people.

#### Musculo-skeletal symptom

James et al. [25] found that reporting of emergency department encounters for spondylosis, intervertebral disc disorders, and other back problems, was less for DHH ASL users (n=19) in comparison with DHH English speakers (n=56) and hearing English speakers.

#### Pregnancy, childbirth or the puerperium

Of the 32 encounters recorded in emergency department records for other complications of pregnancy James et al. [25], only 3 were DHH ASL users compared with 25 DHH English and 4 hearing English speakers.

#### Headache

A secondary data study by Wahlqvist et al. [27] reported that Deaf people with USH1 (n=60) expressed significantly more prevalent problems with headaches compared to the cross section of the Swedish population including those with and without visual difficulties (n=5738) (40% vs 26% respectively).

#### Chronic comorbidity

Kushalnagar et al. [20] reported that the hearing sample has more individuals with comorbidities compared to the Deaf sample (40.5% vs 34.2%), although the hearing sample was older than the Deaf sample which could explain the higher prevalence in the hearing sample.

#### Suicide attempts / suicidal thoughts

The prevalence of suicide attempts in the past year is higher in the Deaf population (2.2%) in the US than observed in the general population (0.4%) [9] and Deaf people reported more suicide attempts in the past year compared with the general population (1.5% vs 0.5%) [28]. Deaf people with Usher Syndrome Type 1 (USH1) are more likely to attempt suicide compared with the general population (16% vs 4%) [27]. James et al., [25] in a study of emergency department records report no suicide ideation and intentional self-inflicted injury reported for DHH ASL users, in comparison with two were reported for DHH English speakers and three for hearing English speakers [25].

#### Health status

Health status was found to be poorer in the Deaf population compared with the general population in the self-report studies by Rogers et al. [29] (EQ-5D mean index values 0.78 vs 0.84), and Shields et al. [10] (43% vs 17% for depression symptoms). However, Kushalnagar et al. [20], found that hearing people had worse overall health status compared with Deaf people, suggesting that age may be a contributing factor, as the mean age of the hearing sample was significantly older than that of the Deaf adults. The Wahlqvist et al. [27] study reported that the USH1 group have greater problems with fatigue (62% versus 49%), and a loss of confidence (16% versus 6%) compared to the general population.

#### Quality of Life

Fellinger et al. [30] and Henning et al. [31] both reported significantly lower Quality of Life as measured by WHOQOL-BREF in Deaf people compared with general populations. The use of the sign language version of WHOQOL-BREF was not reported in Fellinger et al. [30] study.

### Factors identified as influencing health outcomes within the Deaf population

#### LGBTQ+ status

Kushalnagar et al. [32] found that the Deaf LGBTQ population in the US, in comparison with the Deaf non-LGBTQ population, are more likely to have a personal cancer history (24.1% vs 15.2%), more likely to have a lung condition (23.4% vs 15%), and significantly more likely to experience depression/anxiety (33.3% vs 17.9%). Deaf LGBTQ status was also significantly associated with increased risk for arthritis (RR=1.26) and for chronic comorbidity (2 or more medical conditions) (RR=1.25) [32] in comparison with the Deaf non-LGBTQ population. A small-scale study (n=36) reported that the LGBTQ status were not significantly related to the depression score [33]. In a study involving transgender Deaf communities, it was found depression/anxiety was higher for those with nonbinary identities [34].

#### Educational level

Deaf people with university level education scored higher on psychological well-being compared with other Deaf people [16]. In another study, educational levels were found to be significant in explaining psychological well-being score [35]. A secondary data study found that Deaf people who reported low educational levels were more likely to be at risk for cardiovascular disease compared with Deaf people with a four-year college degree or more (OR=5.76) [36]. Two small-scale self-report studies [37, 38] found that more years of education was significantly associated with higher quality of life for Deaf people.

#### Employment and economic status

Small-scale self-report study found that Deaf people who are not in employment have significantly lower mental well-being compared to those who are in employment (SWEMWBS BSL mean score 21.10 vs 23.40) [23]. Wahlqvist et al. [27] report those with USH1 who are in employment are more likely to attempt suicide compared to the general population who are in employment but those with USH1 who are not in employment the differences in suicidal thoughts are not significant compared to the non-working group in the general population [15]. Income status was reported not to have the presence of cardiovascular disease [36].

#### Ethnicity

Although some studies include race/ethnicity when describing the study samples, few studies have considered the influence of ethnicity on health outcomes. Perrodin-Njoku et al. [17] identified consistently poor health outcomes for Black Deaf adults with regard to diabetes, hypertension, heart condition, lung disease, and cancer, as well as comorbidity. Anderson et al. [31] reported that individuals who identified as a racial/ethnic minority significantly had slightly higher levels of perinatal depression than those who identified as White non-Hispanic. Kushalnagar et al. [20] reported no significant difference in race/ethnicity on depression/anxiety outcomes.

#### Gender/sex

Health outcomes by gender/sex were explored in a few studies. Significantly poorer physical well-being outcomes were reported for Deaf females in the validated sign language version study (n=311) [38]. Poorer well-being / quality of life outcomes for Deaf females compared to Deaf males are found [8, 30, 21, 16, 23]. Kushalnagar et al. [20] higher prevalence of depression/anxiety amongst Deaf females. Deaf men were found to have significantly higher blood pressure (15.9%) compared to Deaf women (7.7%) [8].

#### Language and communication

Using inadequate access to direct child-caregiver communication in childhood as the independent variable, Kushalnagar et al. [39] identified that it increased a person’s risks of having diabetes by 12%, hypertension by 10%, lung disease by 19% and cardiovascular disease by 61% and increased risk for depression/anxiety by 34% compared to those Deaf people who had adequate access to indirect family communication and inclusion [39]. No significant difference in the scores for mood or neurosis were found between those Deaf people who used sign language and deaf people who used spoken language [40].

#### Family history/personal medical history

Using the sign language version of the assessment and when the validation has been examined, Munro et al. [41] reported that a clinical sample had a significantly lower mean score for wellbeing (18.57; SD=9.6) compared with a non-clinical sample (27.04, SD=8.68) on the ORS-Auslan. Overall health status was found to be poorer for Deaf people with depression compared to those with no psychological distress or depression [10]. Rogers et al. [42] and Belk et al. [43] found that severity of depression and anxiety was worse for those who self-reported as having mental health difficulties compared to those who did not.

#### Age

It was reported that diagnosis of depression/anxiety was likely to be young in the large-scale study [20], however the age was reported not to have impact on depression outcomes in Deaf populations in the study by Duarte et al. [38].

## DISCUSSION

The findings from this systematic review demonstrate that, in general, physical health and mental health outcomes in Deaf signing populations are worse when compared with general population samples. Additionally, the impact of a health condition on other health outcomes can created further health inequalities, for example, although not a comparison to the general population, Barnett et al. [44] study which involved a whole sample who were overweight/obese (BMI of 25 or greater) and the biometric outcomes were recorded by a research nurse, 13.5% had diabetes, 37.5% had high blood pressure, 53.8% had high cholesterol, 2.9% had heart disease, 39.6% had a PHQ-9 score indicative of at least mild depression. However, the strength and quality of the evidence available is questionable.

Firstly, sample definition is poor with inconsistencies in reporting which add to the difficulties in collating and appraising data concerning health outcomes for Deaf adults. The main issues include inaccurate or imprecise descriptions of participants meaning it is hard to discern in some studies who are Deaf sign language users and some study populations incorporated children and young people without any clear distinction from adults in data subsets. Secondly, some studies do not report whether the health outcomes measured used validated standard instruments in sign language nor report potential issues associated with interpreter-mediated assessment and engagement, particularly with regards to self-reported health data. Thirdly, secondary data analysis comparisons with ‘general population’ data will include some participants who are deaf but not sign language users unless matched ‘hearing’ samples have been constructed. Fourthly, creating binary comparisons between Deaf sign language users and hearing/non-signing people can cover up issues of diversity and intersectionality within Deaf communities. Where comparison groups are matched on a range of demographic variables, these may still hide different circumstances associated with variables e.g. social determinants that are more prevalent amongst Deaf populations e.g. under-employment or direct discrimination.

Furthermore, gaps remain in the knowledge of specific health outcomes as there is no reported health outcome data for the Deaf population in 11 out of the 26 (42.3%) of the ICD-11 disease classification categories, including, for example, diseases of the immune system, visual system and nervous system which indicates clear deficits in health outcome data for this population. The bias towards studies concerning mental health might be in part explained by the longstanding development of specialist mental health services for deaf people in some countries such as the UK and US garnering funding for evidence-based practices. The major neglect of data on physical health outcomes might be related to the considerable difficulties in recording and extracting *routine* health data that is specific enough to differentiate Deaf people from anyone who is categorised with a hearing disability in routine health data collection [45]. For example James et al. [25] in a study on emergency department encounters, highlighted the possibility that DHH ASL users were being mis-recorded as DHH English speakers. The invisibility of the Deaf population within clinical records is likely to contribute to a lack of focus on whether their outcomes are similar to those of the bigger population of adults with a hearing loss or disability but who are not members of a cultural-linguistic minority whose engagement with health services is fundamentally mediated by problems of linguistic access and cultural competence [46]. In addition, the overwhelming majority of the included studies concern Deaf people who reside in economically well-resourced countries. Yet, nearly 80% of people who experience deafness, whether sign language users or not, reside in low- and middle-income countries [1].

The reasons for the health inequalities experienced by Deaf individuals are multiple and complex, both access to and communication with health services and clinicians are commonly cited problems [7]. Around 5% of deaf children have one or more parents who are d/Deaf, meaning that the vast majority are born to hearing parents, who usually have little experience of deafness and often have little or no knowledge of signed languages [47]. Age-appropriate literacy remains a key barrier to accessing information for a great many d/Deaf people and is especially apparent amongst sign language users of previous generations whose access to and quality of education has been particularly poor [48, 49]. The responsiveness of health services and health interventions to provide and promote understanding of health conditions in a signed language is also identified as inadequate in many countries. Deaf individuals are up to 7 times more likely to experience poor health literacy than their hearing counterparts, something which is closely tied to unhealthy behaviours, limited healthcare seeking behaviours, decreased service use and poorer health outcomes [50, 51, 52]. Studies show that Deaf people have limited knowledge of the symptoms of certain medical emergencies, such as heart attacks and strokes, and that in the US, only 61% would contact the emergency services in such cases [53]. Research has also explored the issue of inadequate adaptation of clinical and psychological assessment tools for use with Deaf patients [54, 55], resulting in both under and overdiagnosis of potentially serious health conditions and inadequate tracking of recovery [43, 56]. Understandably, Deaf populations have previously reported feelings of mistrust towards healthcare professionals [7], these populations are also found to be less likely to see the value in healthcare consultations when compared with the general population [57]. Aggravating this, many Deaf patients also have difficulty complaining about the healthcare barriers they face, as complaints processes often do not accommodate for sign language users [58].

Consequentially, healthcare professionals are unaware of the relevant issues, and no action is taken to amend them.

## CONCLUSION

This comprehensive systematic review on health outcomes in Deaf signing populations has highlighted health inequalities in comparison to general populations and within their own communities.

## Author contributions

Contributor role of the following authors are as follows:

**KR** Conceptualization; Data Curation; Formal Analysis; Funding Acquisition; Investigation; Methodology; Project Administration; Resources; Validation; Visualization; Writing – Original Draft Preparation; Writing – Review & Editing

**AR** Data Curation; Formal Analysis; Investigation; Methodology; Project Administration; Resources; Validation; Visualization; Writing – Original Draft Preparation; Writing – Review & Editing

**JH** Investigation; Validation; Writing – Original Draft Preparation; Writing – Review & Editing

**GS** Conceptualization; Formal Analysis; Methodology; Validation; Writing – Original Draft Preparation; Writing – Review & Editing

**AY** Conceptualization; Investigation; Methodology; Validation; Visualization; Writing – Original Draft Preparation; Writing – Review & Editing

## Data Availability

Not applicable as this is a systematic review paper.

## REFERENCES

[1] World Health Organization. Deafness and hearing loss. [Internet]. 2021 [updated 2023 February 27; cited 2023 June 26]. Available from: https://www.who.int/news-room/fact-sheets/detail/deafness-and-hearing-loss.

[2] National Geographic. [Internet]. 2023 [updated 2023 October 19; cited 2023 October 31]. Available from: https://education.nationalgeographic.org/resource/sign-language/

[3] United Nations. [Internet]. 2023 [cited 2023 October 31]. Available from: https://www.un.org/en/observances/sign-languages-day

[4] Sutton-Spence R, Woll B. The linguistics of British Sign Language: an introduction. Cambridge, UK; 1999.

[5] Ladd P. Understanding Deaf Culture: In search of Deafhood. Clevedon; 2003.

[6] Alexander A, Ladd P, Powell S. Deafness might damage your health. Lancet. 2012, Mar 17; 379: (9820):979–81. doi: 10.1016/S0140-6736(11)61670-X.

[7] Emond A, Ridd M, Sutherland H, Allsop L, Alexander A, Kyle J. Access to primary care affects the health of Deaf people. British Journal of General Practice. 2015; 65(631):95–96. doi: 10.3399/bjgp15X683629

[8] Emond A, Ridd M, Sutherland H, Allsop L, Alexander A, Kyle J. The current health of the signing Deaf community in the UK compared with the general population: a cross-sectional study. BMJ Open. 2015, Jan 15; 5:e006668. doi: 10.1136/BMJOPEN-2014-006668.

[9] Barnett S, Klein JD, Pollard PQ, Samar V, Schlehofer D, Starr M, Sutter E, Yang H, Pearson TA. Community Participatory Research With Deaf Sign Language Users to Identify Health Inequities. American Journal of Public Health. 2011 Dec;101(12):2235–8. doi: 10.2105/AJPH.2011.300247.

[10] Shields GE, Rogers KD, Youn, A, Dedotsi S, Davies LM. Health State Values of Deaf British Sign Language (BSL) Users in the UK: An Application of the BSL Version of the EQ-5D-5L. Applied Health Economics and Health Policy. 2020, Jan 16;18:547–556. doi: 10.1007/s40258-019-00546-8

[11] Humphries T, Kushalnagar P, Mathur G, Napoli DJ, Padden C, Rathmann C, Smith C. Avoiding Linguistic Neglect of Deaf Children. Social Service Review. 2016, Dec; 90(4):589–619. doi: 10.1086/689543.

[12] Schenkel LS, Rothman-Marshall G, Schlehofer DA, Towne TL, Burnash DL, Priddy BM. Child maltreatment and trauma exposure among deaf and hard of hearing young adults. Child Abuse & Neglect. 2014, Oct;38(10):1581–9. doi: 10.1016/J.CHIABU.2014.04.010.

[13] Earis H, Reynolds S. Deaf and hard-of-hearing people’s access to primary health care services in North East Essex: A report for North East Essex Primary Care Trust. [Internet] 2009. [cited 2023 January 18]. Available from: http://www.sally-reynolds.com/wp-content/uploads/2012/12/Report-on-access-for-NE-Essex-PCT.pdf

[14] Tranebjærg L, Samson RA, Green GE. Jervell and Lange-Nielsen Syndrome. In: Adam MP, Everman DB, Mirzaa GM, et al., editors. GeneReviews® [Internet]. Seattle (WA): University of Washington, Seattle; 1993-2023. Available from: https://www.ncbi.nlm.nih.gov/books/NBK1405/

[15] Ehn M, Wahlqvist M, Danermark B, Dahlström Ö, Möller C. Health, work, social trust, and financial situation in persons with Usher syndrome type 1. Work. 2018;60(2):209–220. doi: 10.3233/WOR-182731.

[16] Peñacoba C, Garvi D, Gómez L, Álvarez A. Psychological Well-Being, Emotional Intelligence, and Emotional Symptoms in Deaf Adults. American Annals of the Deaf. 2020;165(4):436–452. doi: 10.1353/aad.2020.0029.

[17] Perrodin-Njoku E, Corbett C, Moges-Riedel R, Simms L, Kushalnagar P. Health disparities among Black deaf and hard of hearing Americans as compared to Black hearing Americans: A descriptive cross-sectional study. Medicine. 2022 Jan 14;101(2):e28464. doi: 10.1097/MD.0000000000028464.

[18] Druel V, Hayet H, Esman L, Clavel M, Rougé Bugat ME. Assessment of cancers’ diagnostic stage in a Deaf community - survey about 4363 Deaf patients recorded in French units. BMC Cancer. 2018 Jan 23;18(1):93. doi: 10.1186/s12885-017-3972-3.

[19] James TG, McKee MM, Sullivan MK, Ashton G, Hardy SJ, Santiago Y, Phillips DG, Cheong J. Community-Engaged Needs Assessment of Deaf American Sign Language Users in Florida, 2018. Public Health Reports. 2022 Jul-Aug;137(4):730–738. doi: 10.1177/00333549211026782.

[20] Kushalnagar P, Reesman J, Holcomb T, Ryan C. Prevalence of Anxiety or Depression Diagnosis in Deaf Adults. Journal of Deaf Studies and Deaf Education. 2019 Oct 1;24(4):378–385. doi: 10.1093/deafed/enz017.

[21] Kvam MH, Loeb M, Tambs K. Mental health in deaf adults: symptoms of anxiety and depression among hearing and deaf individuals. Journal of Deaf Studies and Deaf Education. 2007 Winter;12(1):1–7. doi: 10.1093/deafed/enl015.

[22] Werngren-Elgström M, Dehlin O, Iwarsson S. Aspects of quality of life in persons with pre-lingual deafness using sign language: subjective wellbeing, ill-health symptoms, depression and insomnia. Archives of Gerontology and Geriatrics. 2003 Jul-Aug;37(1):13–24. doi: 10.1016/s0167-4943(03)00003-7.

[23] Rogers KD, Dodds C, Campbell M, Young A. The validation of the Short Warwick-Edinburgh Mental Well-Being Scale (SWEMWBS) with deaf British sign language users in the UK. BMC Health and Quality of Life Outcomes. 2018 Jul 24;16(1):145. doi: 10.1186/s12955-018-0976-x.

[24] Simons AN, Moreland CJ, Kushalnagar P. Prevalence of Self-Reported Hypertension in Deaf Adults Who Use American Sign Language. American Journal of Hypertension. 2018 Oct 15;31(11):1215–1220. doi: 10.1093/ajh/hpy111.

[25] James TG, McKee MM, Miller MD, Sullivan MK, Coady KA, Varnes JR, Pearson TA, Yurasek AM, Cheong J. Emergency department utilization among deaf and hard-of-hearing patients: A retrospective chart review. Disability and Health Journal. 2022 Jul;15(3):101327. doi: 10.1016/j.dhjo.2022.101327.

[26] Vichayanrat T, Kositpumivate W. Oral health conditions and behaviors among hearing impaired and normal hearing college students at Ratchasuda College, Nakhon Pathom, Thailand. Southeast Asian Journal of Tropical Medicine and Public Health. 2014 Sep;45(5):1228–35.

[27] Wahlqvist M, Möller K, Möller C, Danermark B. Physical and psychological health, social trust, and financial situation for persons with Usher syndrome type 1. British Journal of Visual Impairment. 2016;34(1):15–25. doi: 10.1177/0264619615610158

[28] Barnett SL, Matthews KA, Sutter EJ, DeWindt LA, Pransky JA, O’Hearn AM, David TM, Pollard RQ, Samar VJ, Pearson TA. Collaboration With Deaf Communities to Conduct Accessible Health Surveillance. American Journal of Preventive Medicine. 2017 Mar;52(3 Suppl 3):S250–S254. doi: 10.1016/j.amepre.2016.10.011.

[29] Rogers KD, Pilling M, Davies L, Belk R, Nassimi-Green C, Young A. Translation, validity and reliability of the British Sign Language (BSL) version of the EQ-5D-5L. Quality of Life Research. 2016 Jul;25(7):1825–34. doi: 10.1007/s11136-016-1235-4.

[30] Fellinger J, Holzinger D, Dobner U, Gerich J, Lehner R, Lenz G, Goldberg D. Mental distress and quality of life in a deaf population. Social Psychiatry and Psychiatric Epidemiology. 2005 Sep;40(9):737–42. doi: 10.1007/s00127-005-0936-8.

[31] Henning MA, Krägeloh CU, Sameshima S, Shepherd D, Shepherd G, Billington R. Access to New Zealand Sign Language interpreters and quality of life for the deaf: a pilot study. Disability and Rehabilitation. 2011;33(25-26):2559–66. doi: 10.3109/09638288.2011.579225.

[32] Kushalnagar P, Miller CA. Health Disparities Among Mid-to-Older Deaf LGBTQ Adults Compared with Mid-to-Older Deaf Non-LGBTQ Adults in the United States. Health Equity. 2019 Oct 30;3(1):541–547. doi: 10.1089/heq.2019.0009.

[33] Anderson ML, Kelly S, Wolf Craig KS, Hostovsky S, Bligh M, Bramande E, Walker K, Biebel K, Byatt N. Creating the Capacity to Screen Deaf Women for Perinatal Depression: A Pilot Study. Midwifery. 2021 Jan; 92:102867. doi: 10.1016/j.midw.2020.102867

[34] Sanfacon K, Leffers A, Miller C, Stabbe O, DeWindt L, Wagner K, Kushalnagar P. Cross-Sectional Analysis of Medical Conditions in the U.S. Deaf Transgender Community. Transgender Health. 2021 Jun 2;6(3):132–138. doi: 10.1089/trgh.2020.0028.

[35] Chapman M, Dammeyer J. The Significance of Deaf Identity for Psychological Well-Being. Journal of Deaf Studies and Deaf Education. 2017 Apr;22(2):187–194. doi: 10.1093/deafed/enw073.

[36] McKee MM, McKee K, Winters P, Sutter E, Pearson T. Higher educational attainment but not higher income is protective for cardiovascular risk in Deaf American Sign Language (ASL) users. Disability and Health Journal. 2014 Jan;7(1):49–55. doi: 10.1016/j.dhjo.2013.07.001.

[37] Ammons D, Engelman A, Kushalnagar P. Quality of Life and Needs of Deaf Informal Caregivers of Loved Ones with Alzheimer’s and Related Dementia. Gerontology & Geriatric Medicine. 2020 Oct 21;6, 2333721420966518. doi: 10.1177/2333721420966518

[38] Duarte SBR, Chaveiro N, de Freitas AR, Barbosa MA, Camey S, Fleck MP, Porto CC, Rodrigues CL, Rodríguez-Martín D. Validation of the WHOQOL-Bref instrument in Brazilian sign language (Libras). Quality of Life Research. 2021 Jan;30(1):303–313. doi: 10.1007/s11136-020-02611-5.

[39] Kushalnagar P, Ryan C, Paludneviciene R, Spellun A, Gulati S. Adverse Childhood Communication Experiences Associated With an Increased Risk of Chronic Diseases in Adults Who Are Deaf. American Journal of Preventive Medicine 2020 Oct;59(4):548–554. doi: 10.1016/j.amepre.2020.04.016.

[40] Øhre B, Volden M, Falkum E, von Tetzchner S. Mental Disorders in Deaf and Hard of Hearing Adult Outpatients: A Comparison of Linguistic Subgroups. Journal of Deaf Studies and Deaf Education. 2017 Jan;22(1):105–117. doi: 10.1093/deafed/enw061.

[41] Munro L, Rodwell J. Validation of an Australian sign language instrument of outcome measurement for adults in mental health settings. Australian and New Zealand Journal of Psychiatry. 2009 Apr;43(4):332–9. doi: 10.1080/00048670902721111.

[42] Rogers KD, Young A, Lovell K, Campbell M, Scott PR, Kendal S. The British Sign Language versions of the Patient Health Questionnaire, the Generalized Anxiety Disorder 7-item Scale, and the Work and Social Adjustment Scale. Journal of Deaf Studies and Deaf Education. 2013 Jan;18(1):110–22. doi: 10.1093/deafed/ens040.

[43] Belk RA, Pilling M, Rogers KD, Lovell K, Young A. The theoretical and practical determination of clinical cut-offs for the British Sign Language versions of PHQ-9 and GAD-7. BMC Psychiatry. 2016 Nov 3;16(1):372. doi: 10.1186/s12888-016-1078-0.

[44] Barnett S, Matthews K, DeWindt L, Sutter E, Samuel-Hodge C, Yang H, Pearson TA. Deaf Weight Wise: A novel randomized clinical trial with Deaf sign language users. Obesity. 2023 Mar 8:,31(4):965–976. doi: 10.1002/oby.23702

[45] Office for National Statistics (ONS). Coronavirus (COVID-19) related deaths by hearing and vision impairment status, England methodology. [Internet]. 2022 [cited 2023 January 18] Available from: https://www.ons.gov.uk/peoplepopulationandcommunity/healthandsocialcare/disability/methodologies/coronaviruscovid19relateddeathsbyhearingandvisionimpairmentstatusenglandmethodology

[46] Hulme C, Rogers K, Young A, Munro K. Deaf Sign Language Users and Audiology Services: A scoping review on cultural competence practices. In: Hulme, C. Improving Patient Experience, Service access and outcomes in NHS Hearing Aid Services for Deaf adults who use British Sign Language. PhD thesis, The University of Manchester; 2022.

[47] Mitchell RE, Karchmer MA. Chasing the Mythical Ten Percent: Parental Hearing Status of Deaf and Hard of Hearing Students in the United States. Sign Language Studies. 2004, Winter; 4(2):138–163. doi: 10.1353/sls.2004.0005

[48] Conrad R. The deaf schoolchild: Language and cognitive function: R. Conrad. London; 1979.

[49] Mayer C. What Really Matters in the Early Literacy Development of Deaf Children. Journal of Deaf Studies and Deaf Education. 2007, June; 12(4):411–431. doi: 10.1093/deafed/enm020.

[50] McKee MM, Paasche-Orlow M, Winters PC, Fiscella K, Zazove P, Sen A, Pearson T. Assessing Health Literacy in Deaf American Sign Language Users. Journal of Health Communication. 2015, Oct;20(2):92–100. doi: 10.1080/10810730.2015.1066468.

[51] Protheroe J, Nutbeam D, Rowlands G. Health literacy: a necessity for increasing participation in health care. British Journal of General Practice. 2009, Oct 1;59(567):721–3. doi: 10.3399/BJGP09X472584.

[52] Naseribooriabadi T, Sadoughi F, Sheikhtaheri A. Barriers and Facilitators of Health Literacy among D/deaf Individuals: A Review Article. Iranian Journal of Public Health. 2017 Nov;46(11):1465–1474.

[53] Margellos-Anast H, Estarziau M, Kaufman G. Cardiovascular disease knowledge among culturally Deaf patients in Chicago. Preventive Medicine. 2006, Mar;42(3):235–239. doi: 10.1016/J.YPMED.2005.12.012.

[54] Palese A, Salvador L, Cozzi D. One-Dimensional Scales for Pain Evaluation Adopted in Italian Nursing Practice: Giving Preference to Deaf Patients. Journal of Nursing Measurement. 2011, Jan;19(2):91–104. doi: 10.1891/1061-3749.19.2.91.

[55] Rogers KD, Ferguson-Coleman E, Young A. Challenges of Realising Patient-Centred Outcomes for Deaf Patients. The Patient: Patient-Centered Outcomes Research. 2018, Feb; 11(1):9–16. doi: 10.1007/s40271-017-0260-x

[56] Young A, Ferguson-Coleman E, Wright B, LeCouteur A. Parental Conceptualizations of Autism and Deafness in British Deaf Children. Journal of Deaf Studies and Deaf Education. 2019, July; 24(3):280–288. doi: 10.1093/deafed/enz002

[57] Sahota O, Boff A, Malthouse K, Qureshi M, Shawcross V. Access to health services for deaf people. London Assembly. [Internet]. 2015 [cited 2023 January 18] Available from: https://www.london.gov.uk/sites/default/files/london_assembly_health_committee_-_access_to_health_services_for_deaf_people_-_june_2015_-_updated.pdf

[58] NHS Liverpool CCG. Understanding Experiences of D/deaf People and People with Hearing Loss in Getting Information and Communication Support from the NHS in Liverpool. 2^nd^ May 2018 Engagement Meeting Report. NHS Liverpool CCGs Engagement Team. [Internet. 2018 [cited 2023, January 18] Available from: https://www.liverpoolccg.nhs.uk/media/3391/liverpool-nhs-final-report-and-actions-deaf-hearing-loss-final.pdf

[59] Crowe TV, Gimire B, Trollo S. The mental health needs of deaf adults in Nepal. International Social Work. 2016; 59(4),508–522. doi: 10.1177/0020872814539983

[60] Horton HK. Linguistic ability and mental health outcomes among deaf people with schizophrenia. Journal of Nervous and Mental Disease. 2010 Sep;198(9):634–42. doi: 10.1097/NMD.0b013e3181e9dd23.

